# Pre exposure hydroxychloroquine prophylaxis for covid-19 in healthcare workers: a retrospective cohort

**DOI:** 10.1101/2020.06.09.20116806

**Authors:** Raja Bhattacharya, Sampurna Chowdhury, Rishav Mukherjee, Anita Nandi, Manish Kulshrestha, Rohini Ghosh, Souvik Saha

**Affiliations:** Associate Professor, Department of General Medicine, Medical College, Kolkata; Post Graduate Trainee, Department of General Medicine, Medical College, Kolkata; Assistant Professor, Department of Microbiology, Medical College, Kolkata; Intern, Medical College, Kolkata; MSc Statistics, Department of Statistics, Presidency University

**Keywords:** Covid19, hydroxychloroquine, pre-exposure prophylaxis, healthcare workers

## Abstract

**BACKGROUND OF STUDY:** While several trials are ongoing for treatment of COVID-19, scientific research on chemoprophylaxis is still lacking even though it has potential to flatten the curve allowing us time to complete research on vaccines.

**OBJECTIVES OF THE STUDY:** To explore the potential of HCQ as a pre-exposure prophylaxis for COVID 19 in health care workers in a tertiary care hospital.

**MATERIALS & METHODS:** We have conducted a retrospective cohort study among 106 Health Care Workers (HCW) exposed to COVID-19 patients, at a tertiary care hospital in India where there was an abrupt cluster outbreak within on duty personnel. HCWs who had voluntarily taken hydroxychloroquine (HCQ) prior to exposure were considered one cohort while those who had not were considered to be the Control group. All participants with a verifiable high risk contact history were tested for COVID-19 by RT-PCR.

**RESULTS:** The two cohorts were comparable in terms of age, gender, co-morbidities and exposure. The primary outcome was incidence rates of RT-PCR positive COVID-19 infection amongst HCQ users and Controls. In this retrospective cohort study, 106 healthcare workers were examined of whom 54 were HCQ users and rest were not. The comparative analysis of incidence of infection between the two groups demonstrated that voluntary HCQ usage was associated with lesser likelihood of developing SARS-CoV-2 infection (4 out of 54 HCW), compared to those who were not on it (20 out of 52 HCW), χ^2^ =14.59, p<0.001. None of the HCQ users noted any serious adverse effects.

**CONCLUSIONS:** This study demonstrated that voluntary HCQ consumption as pre-exposure prophylaxis by HCWs is associated with a statistically significant reduction in risk of SARS-CoV-2. These promising findings therefore highlight the need to examine this association in greater detail among a larger sample using Randomised Controlled Trials (RCT).

## INTRODUCTION

SARS-CoV-2 is a positive sense RNA virus of the family *Coronaviridae* and the etiological agent responsible for the novel pneumonia (COVID-19) which triggered an outbreak towards the end of 2019 in Wuhan, China. Since then, there has been an unprecedented spread of the disease and COVID-19 was declared a ‘global pandemic’ by the WHO on March 11, 2020. As of May 24, 2020 it has affected 5.4 million people and caused 344,419 deaths.^1^ In a rapidly evolving situation such as this, we have often had to resort to repurposing old drugs to expedite the process of prevention or treatment of an unfamiliar disease. For treating viral illnesses, often there is a “one bug - one drug” approach which however fails in times of emerging and re-emerging infections. This is especially true during pandemics - when drug discovery races against time. An alternative approach is, to use a broad spectrum antiviral as a pandemic tool.^2^

To this effect, even before this recent crisis, at least 108 Broad Spectrum Antivirals (BSA) had already been identified that had shown activity against 78 viruses.^3^ These BSAs include 4-aminoquinoline compounds (chloroquine, hydroxychloroquine, amodiaquine) amongst others and have been known to prevent viral entry into host cell.^3,4^ In this study we have chosen to focus on Hydroxychloroquine - particularly it’s role, as an antiviral, in prevention of COVID-19. Hydroxychloroquine (HCQ) is a derivative of Chloroquine (CQ), formulated by introducing a hydroxyl group into CQ and was demonstrated to be much less (∼40%) toxic than CQ in animals.^5^ Both share similar chemical structures and mechanisms of action as a weak base and immunomodulator and have exhibited, in vitro, potent antiviral properties against various viruses.^3^

They are both concentrated in organelles with low pH like the lysosomes and endosomes hence are also called lysosomotropic agents.^4^ CQ increase lysosomal pH and prevent its fusion with auto-phagosomes in vitro.^6^ It can also inhibit endosomal acidification, thus preventing viral entry into host cells. ^7^ A proposal to use CQ as a candidate drug for influenza virus was already in place. In fact in Mouse models, CQ and its derivative HCQ had already been used successfully against Avian Influenza.^8^ However this success was unfortunately not replicated in Influenza A or B and RCTs^9^ failed to demonstrate any preventive role in them. In the case of Coronaviruses, for SARS CoV1, CQ and its derivatives were found to show strong antiviral properties in vitro. Apart from interfering with lysosomal and endosomal activities, it also inhibits terminal glycosylation of ACE2 receptor^10^ which is involved in viral entry. ^11,12^ Impaired terminal glycosylation of ACE2 may reduce the binding efficiency between ACE2 on host cells and the SARS-CoV spike protein.^13^ Moreover, Both CQ and HCQ blocked the transport of SARS-CoV-2 from early endosomes to early lysosomes,^14^ which appears to be a requirement to release the viral genome as in the case of SARS-CoV.^15^

Similar to SARS-CoV-1, SARS-CoV-2 also utilises the surface receptor ACE2 ^16^ for cellular entry. In vitro data have also demonstrated that CQ as well as its derivative HCQ are potent inhibitors of SARS-CoV-2.^14^ In spite of it’s in vitro success in inhibiting SARS-CoV-2, much like in the case of SARS-CoV-1 clinical trials have failed to show any benefit of HCQ as a therapy for SARS-CoV-2 infection.^17^

However, data on its Prophylactic role is still incomplete. Noting the in vitro data and theoretical benefits of HCQ usage, ICMR had proposed its prophylactic use for Health Care Workers in India.^18^

This retrospective cohort study explores the usage of HCQ in a tertiary health care centre in India amongst healthcare workers and investigates its prophylactic potential in prevention of COVID-19 infection.

## METHODS

### STUDY DESIGN AND PATIENT SELECTION

This is a retrospective cohort study based on an online survey of 106 health care personnel, who worked at Medical College, Kolkata, a tertiary care teaching hospital in India, dealing with COVID-19 patients and non COVID patients (mixed COVID Facility), in the first two weeks of May, 2020. In the given period, a cluster outbreak of cases amongst HCWs in this hospital had occurred - with about 28 HCW testing positive over a period of two weeks.

Since late March, Indian Council of Medical Research (ICMR)^18^, which is the apex body of medical research in India, has proposed consumption of HCQ for prophylaxis against COVID-19. In accordance with that guideline, some of the HCWs were voluntarily on Pre-exposure HCQ prophylaxis whereas few others were not. After the outbreak was identified, all those who fulfilled the high risk contact criteria were quarantined and tested for COVID-19 between Day 7-Day 14 of suspected exposure as per Ministry of Health and Family Welfare (MoHFW), Government of India, guidelines.^19^ The end results of these tests were recorded and COVID-19 positive health care workers were sent for isolation and treatment to appropriate facilities after proper contact tracing. We conducted an online survey amongst exposed healthcare workers after their test reports were available. To be included in the survey the participant needed to be a Health Care Worker, currently working at the tertiary care centre and on duty at the same centre during the last 1 month preceding the survey, and was tested for COVID-19 by RT-PCR in the same hospital.

Two groups of HCWs were identified from the date of publishing of the ICMR guidelines^18^ for HCQ prophylaxis (March 23, 2020) till the date on which participants were exposed to their first COVID-19 positive patient. During this period those who started taking HCQ prophylaxis were considered as HCQ users and those who did not take HCQ prophylaxis were considered to be the control group. **(FIGURE 1)**

**Figure 1:**
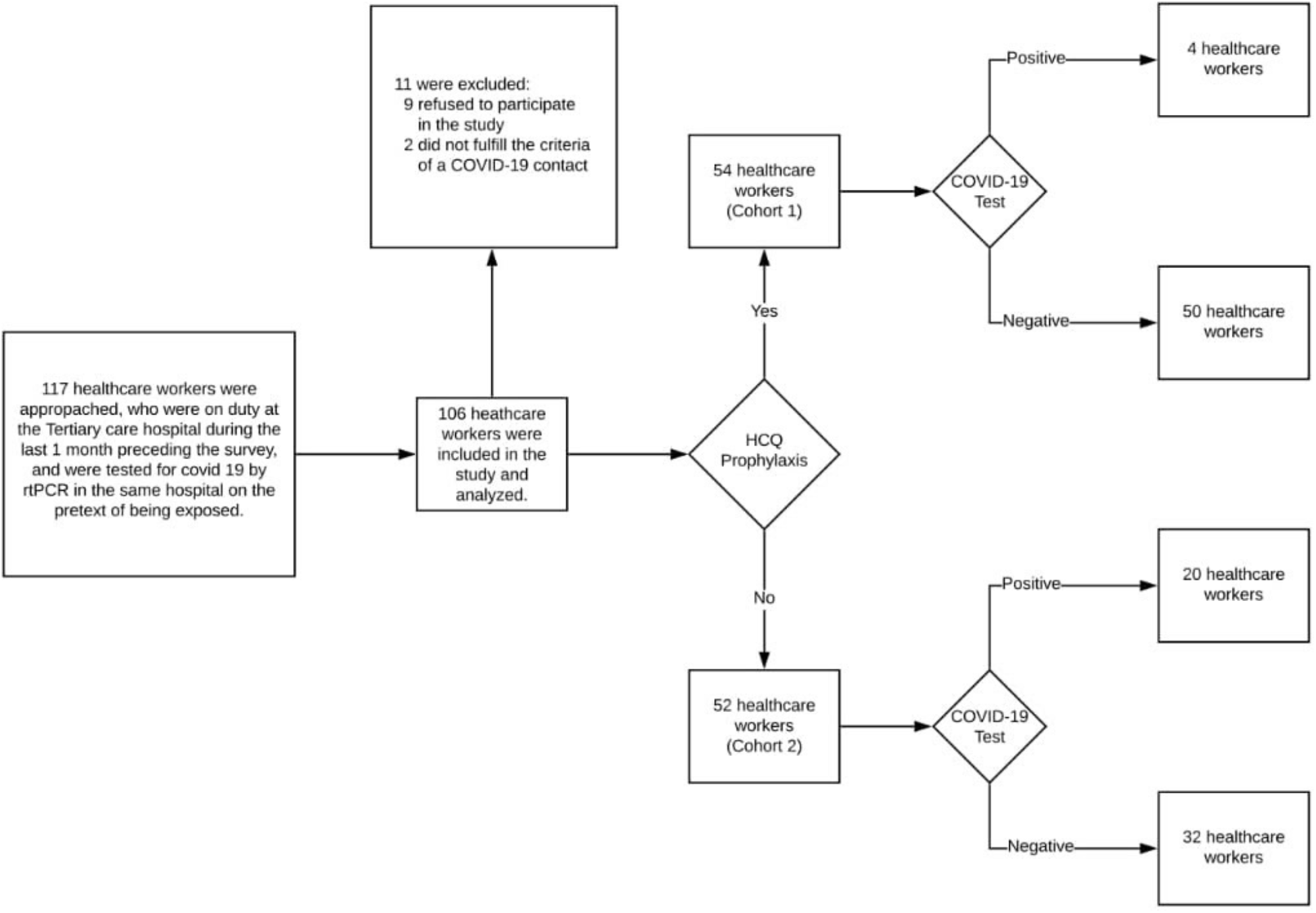
Participant flow chart showing the study cohort.

HCQ users included all the health care workers who were high risk contacts of COVID-19 positive cases & were tested for SARS-CoV-2 Infection in the hospital during the study period, and history of intake of at least the loading dose of hydroxychloroquine prophylaxis and were at any point in their scheduled weekly maintenance dose, as per ICMR guidelines.^18^

Control group included all the health care workers who were high risk contacts of COVID-19 positive cases and were tested for SARS-CoV-2 Infection in the hospital during the same study period, and had either no history of hydroxychloroquine prophylaxis or had history of inadequate intake of HCQ (inadequate loading dose, inadequate schedule maintenance dose or missed doses) as per ICMR guidelines.^18^

The exclusion criteria were refusal to give consent for the study, not being a contact of COVID-19 positive case as per definition given by the National Centre of Disease Control, India (NCDC).^19^ The outcome of Interest was to see whether voluntary HCQ prophylaxis was related to positivity rates by RT- PCR in Health Care Workers.

### TESTING FOR SARS-COV-2 INFECTION

Detection of SARS-CoV-2 in clinical specimens were done by RT-PCR using TaqPath COVID-19 Combo Kit (Applied Biosystems).The real time assay uses the TaqMan fluorogenic probe based chemistry that uses the 5’ nuclease activity of Taq DNA polymerase and enables the detection of a specific PCR product as it accumulates during PCR cycles. COVID-19 Real Time PCR Assay Multiplex-Multiplexed assays that contain three primer/probe sets specific to different SARS-CoV-2 genomic regions and primers/probes for phage MS2 (Internal process control for nucleic acid extraction).^20^ Samples with a result of SARS-CoV-2 Inconclusive were tested again and all were found to be negative and thus, were treated as negative.

### DATA COLLECTION

An internet-based retrospective cohort study was designed where participants volunteered to provide data either on an online form or over the telephone. The online survey form was circulated via online messenger services amongst health care workers who were encouraged to recirculate the same amongst their colleagues. After satisfying the inclusion criteria, a total of 106 participants were considered.

The survey included data on demographic profile like age, sex, presence of co-morbidities (defined as Hypertension, Diabetes, Coronary Artery Disease, Chronic Kidney Disease & COPD),^21,22,23^ and use of hydroxychloroquine prophylaxis as per ICMR guidelines.^18^ Data on type of exposure, nature of contact, use of PPE was also collected in the survey.

### STATISTICAL ANALYSIS

Data was analysed by means of R Software and STATA version 13.1 (StataCorp LP, College Station, TX, USA). Statistical analysis was performed by using test of means of two populations viz. Welch two independent samples T-test, Categorical Data Analysis in the form of contingency tables, partial contingency tables, Chi-squared tests of independence, Chi-squared test of homogeneity, large sample tests of proportion viz. one sample test of proportion and two sample test for proportions. Controlling for possible confounders was done by stratification.

A P value of less than 0.05 was considered statistically significant.

## RESULTS

The two cohort groups of those taking hydroxychloroquine (HCQ) and those not taking hydroxychloroquine (control) were comparable in terms of age, gender, degree of exposure and type of exposure and co-morbidities. **(TABLE 1)**. The mean number of COVID-19 cases with whom the workers had come in contact was also found to be the same in the two groups by a Welch two sample t-test which was found to be 3.

**TABLE 1:** Baseline characteristics of participants in the two cohort groups.

| Parameters | HCQ | Control | p value* |
| --- | --- | --- | --- |
|  | N=54 | N=52 |  |
| Age (mean ± SD) | 26.46 ± 3.93 | 27.71 ± 7.24 | 0.13 |
| Gender |  |  |  |
| Male | 28 (51.85%) | 24 (46.15%) | 0.22 |
| Female | 26 (48.15%) | 28 (53.85%) |  |
| Degree of Exposure |  |  |  |
| Face to face contact | 51 (94.44%) | 41 (78.85%) | 0.85 |
| Direct contact | 29 (53.70%) | 22 (42.31%) | 0.70 |
| Environmental contact | 23 (42.59%) | 19 (36.54%) | 0.28 |
| Type of Exposure |  |  |  |
| Asymptomatic contact | 22 (40.74%) | 19 (36.54%) | 0.48 |
| Symptomatic contact | 32 (59.26%) | 33 (63.46%) |  |
| Comorbidities | 2 (3.70%) | 2 (3.85%) | 0.92 |

Footnote: \*Continuous data was compared using t-test and categorical data was compared using Chi-squared test for homogeneity.

In the HCQ group 4 out of 54 participants were tested to be COVID19 positive, whereas, in the control group 20 out of 52 participants were found to be COVID19 positive. Distribution of HCQ takers and controls across outcome of COVID19 test in univariate analysis indicated the association of risk (Relative Risk = 0.193; 95% CI = 0.071-0.526; p = 0.001) of SARS-CoV-2 infection with lack of pre-exposure HCQ prophylaxis. In this study, it was seen that taking pre-exposure HCQ prophylaxis was associated with an 80.7% reduction in the risk of acquiring a SARS-CoV-2 infection.

Further analysis of incidence of infection between the two groups demonstrated that the incidence of SARS-CoV-2 in those on HCQ pre-exposure prophylaxis was significantly less when compared to the control group with a χ^2^ value of 14.59, p < 0.001.

Adverse effects, mostly mild, i.e not requiring any intervention medical or otherwise, not requiring any hospitalisation, were noted in 29.8% of those on HCQ Prophylaxis **(FIGURE 2)**, thus corroborating previous treatment trials.^24^ Among those taking hydroxychloroquine prophylaxis, GI upset (19.1%), skin rash (6.4%) and headache (4.3%) were reported as adverse effects. GI upset was found to be the most common adverse effect by paired proportion tests. None of the study participants reported discontinuing the drug at the time of survey.

**Figure 2:**
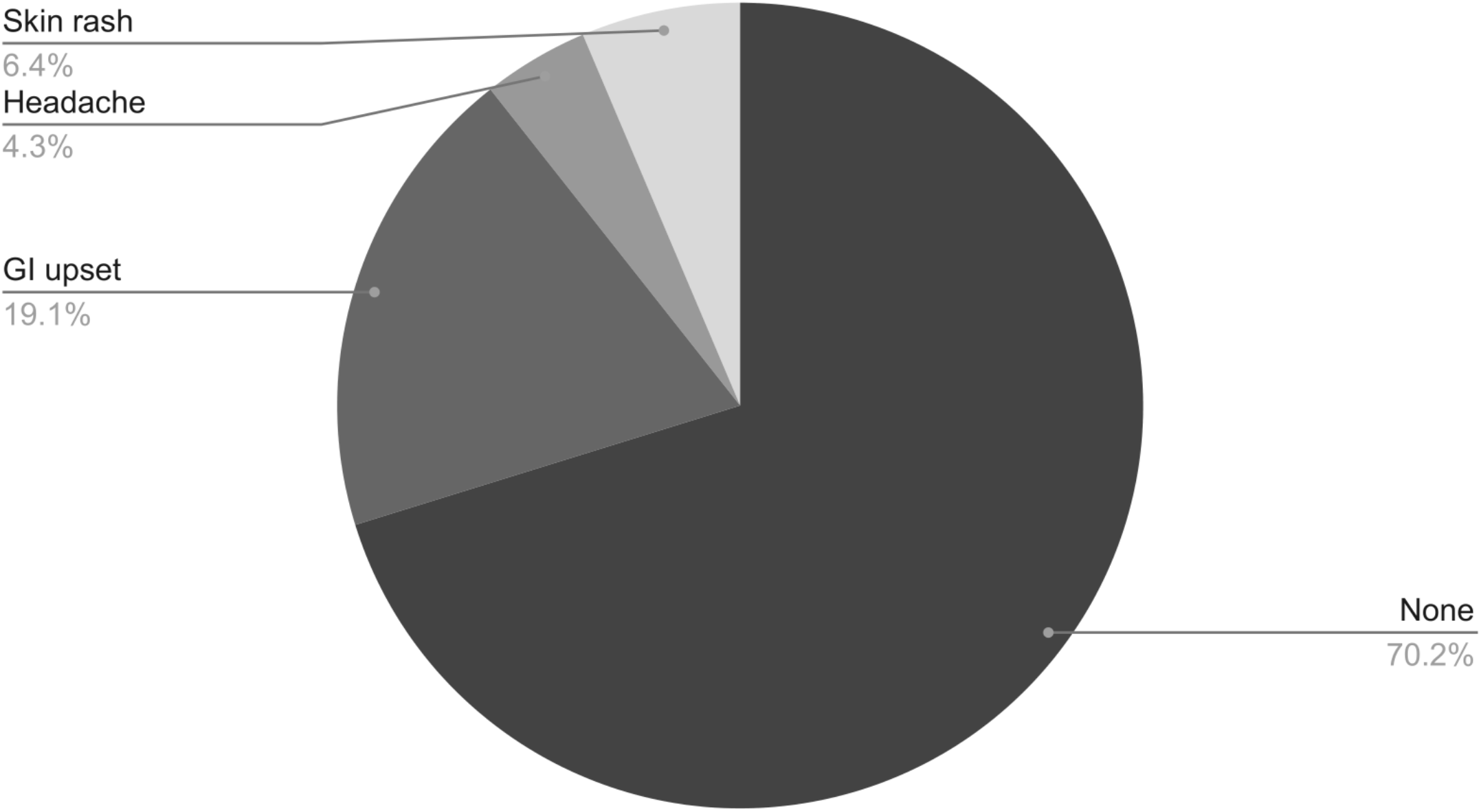
Pie chart showing the adverse effects among those taking HCQ Prophylaxis.

Univariate analysis of distribution of COVID-19 test results in the two cohort groups for different exposures was done. **(TABLE 2, TABLE 3)**. It was also seen that the healthcare workers who were exposed to symptomatic COVID19 positive patients were more likely to develop SARS-CoV-2 infection compared to those exposed to an asymptomatic COVID19 patient (Odds ratio = 6.046; 95% CI = 1.672 to 21.858; p = 0.006).

**TABLE 2:**
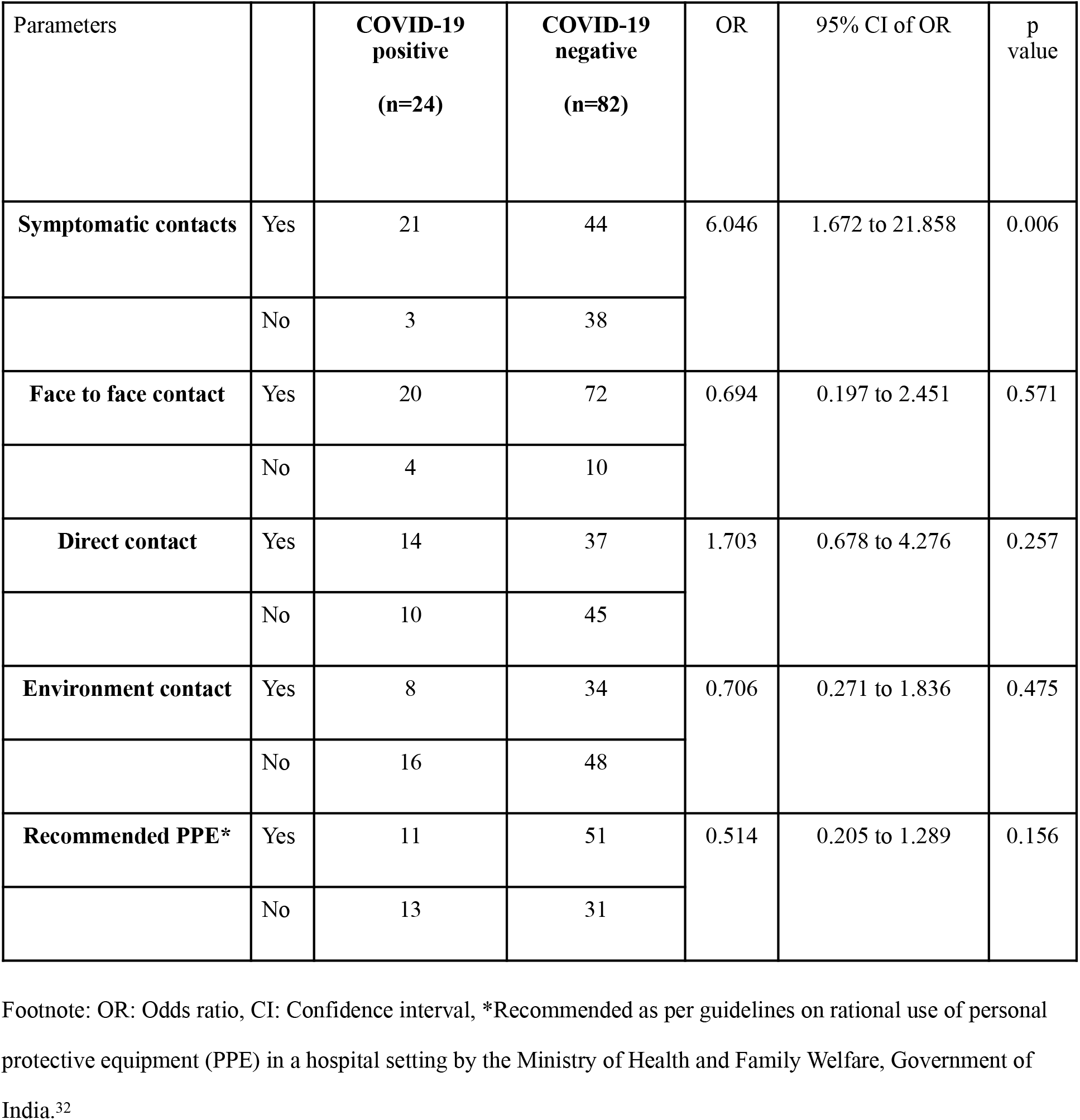
Univariate analysis of distribution of COVID-19 test results for different exposures.

**TABLE 3:** Two way frequency tables showing distribution of COVID-19 test results in the two cohort groups after restricting observations for different parameters

| Parameters |  | **COVID-19**<br>**positive**<br>**(n=24)** | **COVID-19**<br>**negative**<br>**(n=82)** | TOTAL | OR | 95% CI of OR | p<br>value |
| --- | --- | --- | --- | --- | --- | --- | --- |
| **Symptomatic**<br>**contacts** | HCQ+ | 3 (9.38%) | 29 (90.63%) | 32 (100%) | 2.172 | 0.211-22.369 | 0.514 |
|  | HCQ- | 18 (54.55%) | 15 (45.45%) | 33 (100%) | 10.200 | 2.024-51.413 | 0.005 |
| **Face to face**<br>**contact** | HCQ+ | 4 (7.84%) | 47 (92.16%) | 51 (100%) | 0.663 | 0.029-14.972 | 0.796 |
|  | HCQ- | 16 (39.02%) | 25 (60.98%) | 41 (100%) | 1.120 | 0.282-4.450 | 0.872 |
| **Direct contact** | HCQ+ | 3(10.34%) | 26(89.66%) | 29(100%) | 2.769 | 0.269-28.468 | 0.392 |
|  | HCQ- | 11(50.00%) | 11(50.00%) | 22(100%) | 2.333 | 0.743-7.324 | 0.147 |
| **Environment**<br>**contact** | HCQ+ | 0(0.00%) | 23(100%) | 23(100%) | 0.130 | 0.007-2.543 | 0.179 |
|  | HCQ- | 8(42.11%) | 11(57.89%) | 19(100%) | 1.273 | 0.401-4.037 | 0.682 |
| **Recommended**<br>**PPE\*** | HCQ+ | 3(7.69%) | 36(92.31%) | 39(100%) | 1.167 | 0.112-12.183 | 0.898 |
|  | HCQ- | 8(34.78%) | 15(65.22%) | 23(100%) | 0.756 | 0.244-2.345 | 0.628 |

Footnote: OR: Odds ratio, CI: Confidence interval, \*Recommended as per guidelines on rational use of personal protective equipment (PPE) in a hospital setting by the Ministry of Health and Family Welfare, Government of India.<sup>32</sup>

## DISCUSSION

The COVID-19 pandemic has put healthcare systems across the world in crisis with significant social and economic burden on countries. A number of clinical trials^25^ are underway to test the efficacy of several repurposed drugs including chloroquine (CQ), hydroxychloroquine (HCQ), Ivermectin, Remdesivir, Ritonavir/Lopinavir for treatment of COVID-19. None have so far shown exceptional results.^26,24,27^ Of these, although HCQ has gathered particular worldwide attention based on in vitro results that demonstrate efficacy against SARS-CoV-2, there is lack of evidence to suggest that HCQ offers any significant additional clinical benefit for the treatment of hospitalised COVID-19 patients.

Although unprecedented progress has been made amidst this pandemic towards development of vaccines, including three candidate vaccines already in clinical trials, most estimates place the timeline for the launch of a safe and effective vaccine at least more than a year away. This highlights the need for possible alternative vaccines for preventing COVID-19. Chemoprophylaxis has been demonstrated to be a successful modality in preventive medicine in a number of other infectious diseases including malaria, HIV and influenza. A mathematical model^28^ exploring effectiveness of prophylaxis vs treatment in an Influenza pandemic predicted that targeted prophylaxis could flatten the curve by 6-18 months during which effective vaccines against the disease could be developed. However, despite chemoprophylaxis being a promising modality, research on this subject in the context of COVID-19 is currently a missing link. The first in vitro data on prophylaxis against COVID-19 by HCQ or CQ was put forward by Yao et al^29^ In this study, HCQ was found to be more potent than CQ in achieving EC50. Several other in vitro studies^14,30^ found a preventive role for CQ/HCQ. As discussed in the introduction of this article, COVID-19 initiates viral entry through a surface ACE2 receptor, initial viral replication is followed by systemic inflammation. CQ/HCQ can prevent viral entry has been demonstrated by Wang et al^30^ through their time-of-addition studies. Based on these encouraging preclinical trials and supported further by preliminary internal observational studies,^31^ ICMR, which is the key government body handling India’s COVID response, recommended the use of HCQ by high risk individuals such as HCW for prevention of COVID-19. Given the well-established safety profile in rheumatology practice with HCQ being safe even during pregnancy, ICMR’s recommendation is justified, taking into consideration the substantial risks faced by healthcare workers.

The current retrospective cohort study therefore aimed to investigate if hydroxychloroquine could be effective as a pre-exposure prophylaxis for COVID-19 among high risk individuals such as healthcare workers. The results from this study, demonstrate that voluntary consumption of HCQ as prophylaxis among high risk individuals was associated with a significantly reduced risk of testing positive for COVID19. Incidentally, one COVID-19 positive participant who was a long-term user of HCQ for Rheumatoid Arthritis, reported discontinuing the drug 1 month prior to exposure. It was also observed that among HCQ takers 32 were exposed to a symptomatic contact out of which only 3 (9.38%) were positive whereas in the control group 33 were exposed to a symptomatic contact out of which 18 (54.55%) tested to be positive. Similarly, it was seen that proportions of those testing to be positive were less in HCQ takers among those having a face to face contact, direct contact and environmental contact. Those who used recommended^32^ personal protective equipment (PPE), even among them, 3 out of 39 (7.69%) HCQ takers were tested to be positive whereas 8 out of 23 (34.78%) controls acquired the infection. (Table 3). Those exposed to a symptomatic patient were more likely to acquire the infection than those exposed to asymptomatic patients (OR = 6.046; 95% C I= 1.672 to 21.858; p = 0.006). These observations need to be further analysed by future studies. The current study also validated the known safety profile for HCQ with no serious adverse events reported by the participants. This study represents a clinical evidence on the potential role of HCQ as prophylaxis.

Our study design also aimed to include those members in the control group who were taking inadequate dosage of HCQ, as per ICMR guidelines. However, all participants were either taking HCQ as per guidelines or not taking at all. Thus, no data on inadequate HCQ dosage could be analysed. Despite a strong statistical association, a cause-effect relationship can not be inferred. The data included young, healthy individuals with no or limited relevant underlying health conditions, reflecting the demographics of health care workers. This warrants a conservative interpretation of our findings and the statistical significance needs to be explored further. Thus, we recommend further validation through large scale RCT to interrogate the significant association between voluntary HCQ consumption and reduced clinical risk of contracting COVID-19 that emerged from this study.

## LIMITATIONS

Non random sampling was done based on a voluntary response online survey and thus sampling bias and recall bias might be present in the data set. Being a retrospective cohort study it is limited in its ability to measure all confounding factors. Blinding was not done at any level.

At the time this study was carried out, no prevalence data was available in mixed Covid facilities catering to both covid and non-covid patients. Due to an unmet need for data, a sample size calculation was not appropriate. Moreover, transmission of COVID 19 to healthcare workers in this setting was not only possible from patients alone but also from infected coworkers often sharing common areas of the hospital such as lounges, living quarters, dining spaces etc.

A therapeutic dose monitoring for Hydroxychloroquine was not carried out owing to the questionnaire based nature of the study. Study participants were expected to follow Indian Council of Medical Research’s guideline on hydroxychloroquine prophylaxis - however, validation of the dose and its frequency was beyond the scope of the present study.

Among those not taking HCQ a higher number of positives might be because of their risk taking behaviour that prevents them from taking prophylaxis and protective measures as well. On the contrary it might be so that there is a false sense of protection among HCQ takers that predisposes them to more risk-taking behaviour.

The adverse effects which require special assessment like bradycardia and prolonged QT interval could not be recorded and the data was based solely on patient history.

## CONCLUSION

In this retrospective cohort study involving healthcare workers who had been exposed to SARS-CoV-2, voluntary pre-exposure Hydroxychloroquine use was associated with a statistically significant lowered risk of testing positive for SARS-CoV-2. No serious adverse effects were found among those taking HCQ Prophylaxis.

## Data Availability

All relevant data is available within manuscript file.

## ABBREVIATIONS

HCQ: Hydroxychloroquine
CQ: Chloroquine
ICMR: Indian Council Of Medical Research
MOHFW: Ministry of Health & Family Welfare
PPE: Personal Protective Equipment
PCR: Polymerase Chain Reaction

## ACKNOWLEDGEMENT

We acknowledge the dedication, commitment, and sacrifice of the staff and personnel in our institution during the COVID-19 crisis and the suffering and loss of our patients as well as in their families and our community. We would like to extend our gratitude to the Department of Medicine and Department of Microbiology, Medical College, Kolkata for supporting us.

